# Multi-objective Risk-based Resource Allocation for Urban Pandemic Preparedness: The COVID-19 Case in Bogotá, Colombia

**DOI:** 10.1101/2021.02.24.21252407

**Authors:** Jeisson Prieto, Jonatan Gomez

## Abstract

Determining how best to allocate resources to be used during a pandemic is a strategic decision that directly affects the success of pandemic response operations. However, government agencies have finite resources, so they can’t monitor everything all of the time: they have to decide how best to allocate their scarce resources (i.e., budget for antivirals and preventive vaccinations, Intensive Care Unit (ICU), ventilators, non-intensive Care Unit (non-ICU), doctors) across a broad range of risk exposures (i.e., geographic spread, routes of transmission, overall poverty, medical preconditions). This paper establishes a comprehensive risk-based emergency management framework that could be used by decision-makers to determine how best to manage medical resources, as well as suggest patient allocation among hospitals and alternative healthcare facilities. A set of risk indexes are proposed by modeling the randomness and uncertainty of allocating resources in a pandemic. The city understudy is modeled as a Euclidean complex network, where depending on the neighborhood influence of allocating a resource in a demand point (i.e., informing citizens, limit social contact, allocate a new hospital) different network configurations are proposed. Finally, a multi-objective risk-based resource allocation (MoRRA) framework is proposed to optimize the allocation of resources in pandemics. The applicability of the framework is shown by the identification of high-risk areas where to prioritize the resource allocation during the current COVID-19 pandemic in Bogotá, Colombia.

## 1. Introduction

Pandemics are large-scale outbreaks of infectious diseases that cause significant social, political, and economical disruption (PAHO, 2009; Koyuncu and Erol, 2010; Madhav et al., 2017). Policy attention has focused on the need to identify emerging outbreaks that might lead Pandemics, and to expand investment to build preparedness and health capacity (Lederberg et al., 2003). In the preparedness pandemic, effective allocation of limited health resources (i.e., budget for antivirals and preventive vaccinations, Intensive Care Unit (ICU), ventilators, non-intensive Care Unit (non-ICU), doctors) plays a critical role in order to reduce the number of cases, hospitalization, and deaths. Despite the great advances in prevention and treatment of infectious diseases, the world is unaware to respond to Pandemic or any similarly global public-health emergency (Who, 2011; Madhav et al., 2017).

In most countries, health care systems operate at or above maximally designed capacity. Many hospitals just do not have sufficient pre-existing resources to respond to surge capacity in an outbreak (Biddison et al., 2019). Unlike with natural disasters, where the greatest need for resources often occurs early in the time course, pandemic resource requirements will build over months. Outbreaks that become pandemics generally do not take hold in multiple locations at exactly the same time, they are geographically and temporally patchy (Koyuncu and Erol, 2010).

Many government agencies and health planners are responsible for overseeing and monitoring future outbreaks. However, they can’t monitor everything all of the time, they have to decide how best to allocate their scarce resources across a broad range of risk exposures. This is called “risk-based resource allocation.” (Farrell et al., 2013). Different types of government agencies face risk-based resource allocation decisions: agricultural land and water resources (Romero, 2000; Wolgin, 1975; Li et al., 2016), system design in a Distributed Environment (Yeddanapudi et al., 2008; Qiu et al., 2008), terrorism (Willis, 2007; Quadrifoglio, 2008; Ray et al., 2009), and Natural Hazards (i.e., tornados, hurricanes, earthquakes) (Vaziri et al., 2010; Zolfaghari and Peyghaleh, 2015; Murphy and Gardoni, 2007; Vaziri, 2008). In the Risk-based resource allocation for the pandemic response, a demand point has one (or more) associated risk (i.e., geographic spread, routes of transmission, risk factors for infection, overall poverty, medical preconditions) and the objective is to choose the amount to be invested in several interventions which minimize the overall risk exposed by the demand points according to budget constraints and health benefits. Due to the randomness and uncertainty of conditions, not only one but a set of risks may adversely affect the allocation of resources in the geographical space. Then, the objectives (one objective for each risk that a demand point may be exposed) must be optimized simultaneously (Yan and Haimes, 2011; Sun et al., 2014), but there exists a trade-off among objectives, i.e., an improvement gained for one objective is only achieved by making concessions to another objective.

This paper aims to describe and illustrate a Multi-objective Risk-based Resource Allocation framework (MoRRA) that could be used by decision-makers to determine how best to manage medical resources, as well as suggest patient allocation among hospitals and alternative healthcare facilities. In MoRRA, different definitions of risk-based resource allocation are given depending on the geographical space and its neighborhood configuration. This study was carried out during the COVID-19 in Bogotá, Colombia to identify geographic areas with high-risk factors in where to prioritize surveillance to control the outbreak and to generate recommendations for future outbreaks.

The remainder of this paper is organized as follows. In the next section, the background knowledge, the risk-based resource allocation (RRA) problem, and the formulation of the multi-objective RRA (MoRRA) are given. In Section 3, the experimental setup for MoRRA is described. Afterward, numerical results of a case study (the current COVID-19 Pandemic in Bogotá, Colombia) are presented in section 4 where it is demonstrated how MoRRA could help decisionmakers to determine the resource allocation and potential resource shortages in the healthcare system. Finally, conclusions and potential future developments are discussed in the last section.

## 2. Risk-based Resource Allocation in a Multiobjective Framework

Following the risk-based resource allocation methodology proposed in (Farrell et al., 2013), the proposed framework involves three main stages. The first stage is the identification and definition of the risk. The second stage is the estimation of the level of risk posed in a demand point. Once the risk has been defined and measured, an optimal strategy is proposed to minimize the risk exposure.

### 2.1. Risk definition

Although there are different definitions of risk, we use the one given by (Kelman, 2018). Risk is composed by two components, *hazard*, and *vulnerability*.

#### Definition 1.

**Hazard** is the probability that a disaster (i.e., COVID-19) occurs.

#### Definition 2.

**Vulnerability** is the possibility that damages (i.e., fatalities, injuries, property damage, or other consequences) occur at a demand point because a resource is not allocated.

Risk is then defined as the expected damages due to a particular hazard for a given area and reference period. Based on mathematical calculations, the risk of the demand point *i* can be determined as a product of hazard (*H*) and vulnerability (*V*) (DHA, 1992).

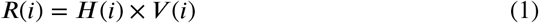

### 2.2. Risk measuring

#### 2.2.1. Hazard assessment

The hazard assessment describes the identification of what hazards can be expected and how they might change in the short and medium-term as a result of environmental phenomena or processes (Kelman, 2018). First of all, all of the potential hazards are identified. Then the areas that could be affected by the hazard are marked, this is called Hazard Mapping. The magnitude, intensity, and frequency of the hazards are determined and the causes of the hazards are investigated. Hazards could include earthquakes, volcanic eruptions, floods, drought, cyclones, and epidemics.

#### 2.2.2. Vulnerability assessment

Vulnerability Assessment describes the degree to which socioeconomic systems and physical assets in geographic areas are either susceptible or resilient to the impact of a disaster (i.e., pandemic). Several models have been proposed to establish vulnerable urban areas over the infectious disease domain, i.e., vector-borne diseases (Hagenlocher et al., 2014), Dengue (de Mattos Almeida et al., 2007), malaria (Kienberger and Hagenlocher, 2014; Hagenlocher and Castro, 2015), Ebola (Moore et al., 2017), and COVID-19 (Mishra et al., 2020; Prieto et al., 2021).

### 2.3. Risk strategy

#### 2.3.1. Resource Allocation Problem

The resource allocation problem seeks to find an optimal allocation of a fixed amount of resources to activities to minimize the cost incurred by the allocation. Given a finite set of resources ℛ = {(*r*_*1*_, *r*_*2*_, … *r*_*a*_) | *r*_i_ ∈ ℝ^+^} whose total amount is equal to *T*, it is required to allocate it to *a* activities so that the objective value *f* (ℛ) is minimized, see equation 2. The objective value may be interpreted as the cost or loss, or the profit or reward, incurred by the resulting allocation (Katoh and Ibaraki, 1998).

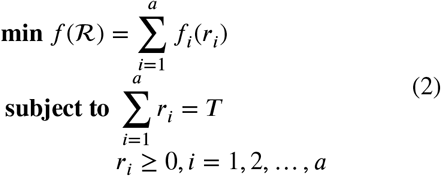

where *r*_*i*_ represents the amount of resource allocated to activity *i* and *f*_*i*_(*r*_*i*_) is the cost incurred by allocate the resource *r*_*i*_ at the *i*-th activity. If the resource is divisible, *r*_*i*_ is a continuous variable that can take any non-negative value. If it represents persons, processors, or trucks, on the other hand, variable *r*_*i*_ becomes a discrete variable that takes non-negative integer values (*discrete resource allocation problem*).

#### 2.3.2. Urban space

Let 𝒮 the geographical space under study (i.e. state, country, or city) defined in terms of a finite set of *P* smaller spatial units (i.e. countries, census tracts, or zip codes); that is 𝒮 = {1, 2, …, *P*}; is an Euclidean complex network *G* = (𝒮, *E*) (Van Der Hofstad, 2017), where the spatial units (or vertex) 𝒮 are located in some position of the 2D Euclidean space and edges *E* are connection between two nodes given the spatial relation *meet*.

##### Definition 3.

The spatial relation *meet*(*i, j*) occurs when *i* has at least one point in common with *j* in the boundary, but their interiors do not intersect (Egenhofer, 1990).

##### Definition 4.

An **adjacent vertex** *u* of a vertex *v* in a graph *G* is a vertex that is connected to *v* by an edge (i.e., *meet*(*u, v*)).

##### Definition 5.

The **neighborhood** of a vertex *v* (*N*(*v*)) in a graph *G* is the subgraph of *G* induced by all vertices adjacent to *v*.

##### Definition 6.

Regardless the metric space under consideration (points, spatial units, binary strings, DNA strands) (Prieto et al., 2019), we will call 𝒩 the **neighborhood class** of all neighborhoods in a graph *G*, i.e., 𝒩 = {*N*(*v*)|*v* ∈ 𝒮}.

#### 2.3.3. Risk-based Resource Allocation Problem (RRA)

Let Λ be a risk values associated for each spatial unit in 𝒮; that is Λ = {(*λ*_1,_ *λ*_2_ … *λ*_p_)| *λ*_*i*_ ∈ ℝ^+^}, the **Risk-based Resource Allocation Problem**, looks for the optimal way to allocate the resources *R* to each demand point (spatial unit) *i* such that the overall risk over 𝒮 is minimized. Here, the cost incurred *f*_Λ,*i*_(*r*_*i*_) by allocate the resource *r*_*i*_ at the *i*-th activity depends on the neighborhood influence of allocate a resource in *i* (i.e., informing citizens, limit social contact, allocate a new hospital), see equation 3.

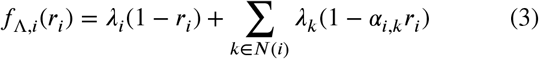

where *r*_*i*_ is the impact factor to allocate a resource to *i* demand point (0 ≤ *r*_*i*_ *≤*1), *N*(*i*) is the neighborhood of *i*-th demand point, and *α*_*i,k*_ is the influence factor in *k* when a resource is allocated in *i* (0 ≤ *α*_*i,k*_ *≤* 1).

Then, the objective function *f*_Λ_ is calculated among spatial units 𝒮.

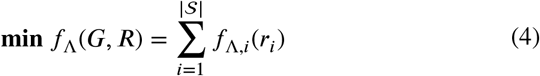

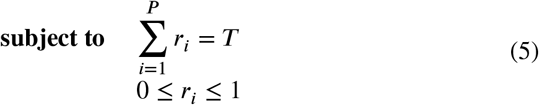

Here, depending on the network configuration, three configurations in RRA are proposed (Fig. 1).

**Fig. 1:**
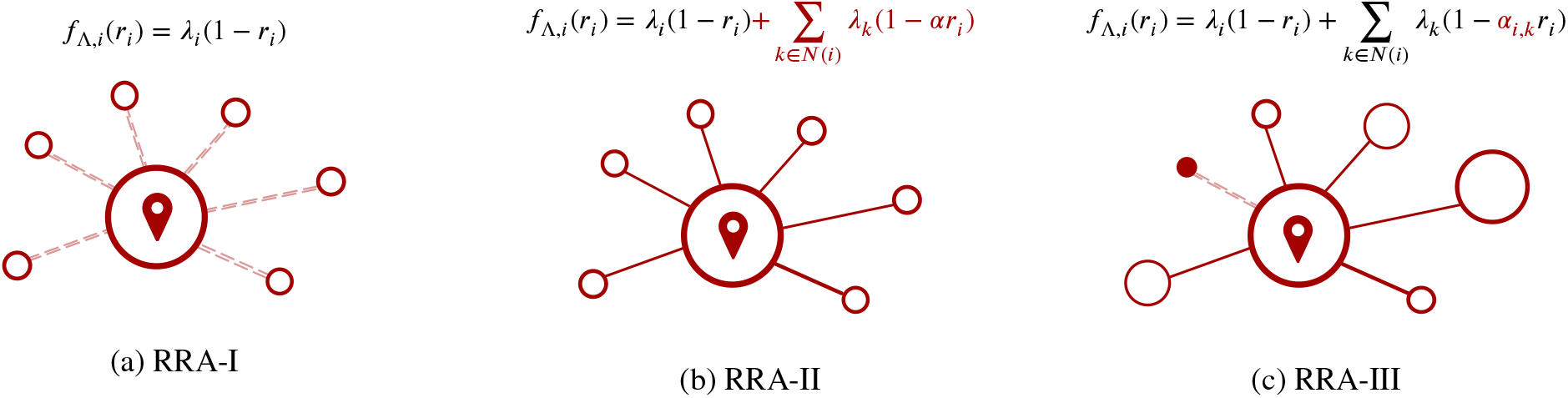
Different configurations of the Risk-based Resource Allocation problem (RRA). Here, the cost incurred *f*_Λ,*i*_(*r*_*i*_) by allocate the resource *r*_*i*_ at the *i*-th activity depends on the neighborhood influence. Without neighbor influence (RRA-I) (left), with neighbor influence at same scale (RRA-II) (middle), and with neighbor influence at different scale (RRA-III) (right).

##### Definition 7.

The **RRA-I** configuration happens when there are not neighborhood influence (∀*i* ∈ 𝒮, *N*(*i*) = Ø). So, the cost incurred *f*_Λ,*i*_(*r*_*i*_) is defined as.

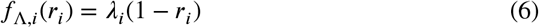

##### Definition 8.

The **RRA-II** configuration happens when there are neighborhood influence at same scale (*α* _*i,k*_ = *a*). So, the cost incurred *f*_Λ,*i*_(*r*_*i*_) is defined as.

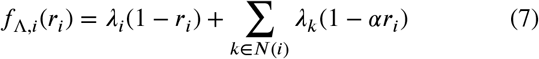

##### Definition 9.

The **RRA-III** configuration happens when there is neighborhood influence at a different scale. So, the cost incurred *f*_Λ,*i*_(*r*_*i*_) is defined as.

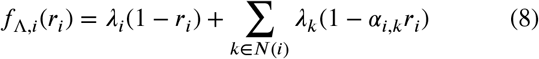

Using the adjacency matrix *A* of *G*, where the *α*_*i,k*_ are the the weight of the edge *w*(*i, k*), the objective function can be evaluated in terms of *A*, see Appendix A.

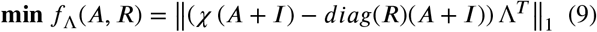

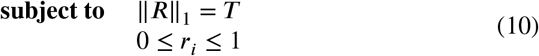

where *χ* is the Indicator function that determines when a value of *A* is different to 0, *I* is the identity matrix, *diag* is the function that diagonalizes the vector of resources *R*, and Λ are the risks associated with the demand points.

#### 2.3.4. Multi-objective RRA (MoRRA)

The **multi-objective optimization problem (MoP)** can be mathematically defined as follows.

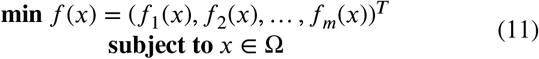

where *x* = (*x*_1_, *x*_2_, …, *x*_*n*_)^*T*^ is the *n*-dimensional decision variable vector from the decision space Ω; *f* : Ω → Θ ℝ^*m*^ consists a set of the *m* objective functions that map *x* from *n*-dimensional decision space Ω to *m*-dimensional objective space Θ.

##### Definition 10.

Given two decision vectors *x, y* ϵ Ω, *x* is said to Pareto **dominate** *y*, denoted by *x ≺ y*, iff *f*_*i*_(*x*) ≤ *f*_*i*_(*y*), for every *i* ∈ {1, 2, …, *m*}, and *f*_*j*_(*x*) *< f*_*j*_(*y*), for at least one index *j* ∈ {1, 2, …, *m*}.

##### Definition 11.

A decision vector *x*^*^ ∈ Ω is **Pareto optimal** iff there is no *x* ∈ Ω such that *x ≺ x*^*^.

##### Definition 12.

The **Pareto set (PS)** is defined as

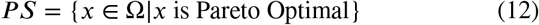

##### Definition 13.

The **Pareto Front (PF)** is defined as

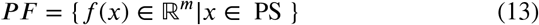

Since objectives in (11) conflicted with each other, no point in Ω simultaneously minimizes all the objectives. The best trade-offs among the objectives can be defined in terms of PF.

Due to the randomness and uncertainty of conditions (environmental, operational), RRA also brings many risks that may adversely affect the allocation of resources in the geographical space. Therefore, it is necessary to introduce a comprehensive set of risk indexes by modeling the randomness and uncertainty of the RRA problems. Then, **Multi-objective Risk-based Resource Allocation** aims to optimal way to allocate *R* to each demand point (spatial unit) *i*, in a set of *M* risk indexes; that is 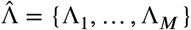.

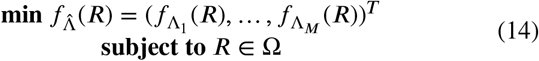

## 3. Resource allocation for COVID-19 in Bogotá, Colombia

### 3.1. Study area and data sources

The applicability of the problem is shown for the curent COVID-19 Pandemic in Bogotá city, the largest and crowded city in Colombia. Bogotá is a metropolitan city with 7.412.566 inhabitants living in an area of 1775km (995km urban and 718km rural), at an altitude of 2640m, with an annual temperature ranging from 6 to 20°C, and annual precipitation of over 840mm. Bogotá is composed of 621 Urban Sectors (Urban Sector is a cartographic the division created by the National Administrative Department of Statistics (DANE) (DANE, 2018a).) Fig. 2 shows the distribution of the Urban sectors over Bogotá.

**Fig. 2:**
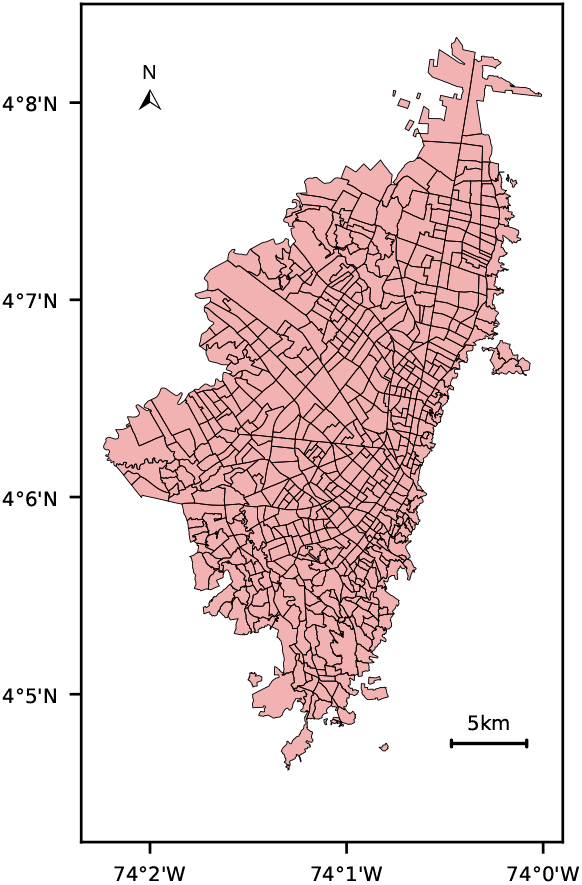
Spatial distribution of Bogotá, Colombia using Urban sectors.

Information was obtained from the National Department of Statistics (DANE), District Planning Secretary of Bogotá (SDP), and the District Mobility Secretary of Bogotá (SDM). Data comprised public information about demographic, transportation, socio-economic, and health conditions reported from 2011 to 2020. A summary of the datasets is presented as follows:

- **MON_2017 (SDP, 2011, 2017):** Dataset provided by SDP containing a set monograph which provides a physical, demographic and socioeconomic description of Bogotá and its districts.
- **SDM_2017 (SDM, 2018):** Dataset provided by SDM presenting detailed official information of mobility characterization in Bogotá.
- **CNPV_2018 (DANE, 2018a)**: Dataset provided by DANE containing the national census made in 2018 which provides socio-demographic statistics of Colombia.
- **DANE_2018 (DANE, 2018b):** Dataset provided by DANE containing the results of the Multidimensional Poverty Index which encompasses educational and health quality, work and housing conditions, and access to public services.
- **DANE_2020 (DANE, 2020):** Dataset provided by DANE presenting a vulnerability index based on demographic and health conditions relevant for COVID-19 pandemic.

The complex network for the different RRA configurations is built (Fig. 3). For RRA-II, the *α* value is fixed in 0.5, which means that influence in the neighborhood is half when a resource is allocated. For RRA-III, the *α*_*ik*_ values are fixed depending on the distance between the spatial units; that is less distance more influence. The *α*_*ik*_ are normalized over the range 0 (less influence) and 0.5 (more influence).

**Fig. 3:**
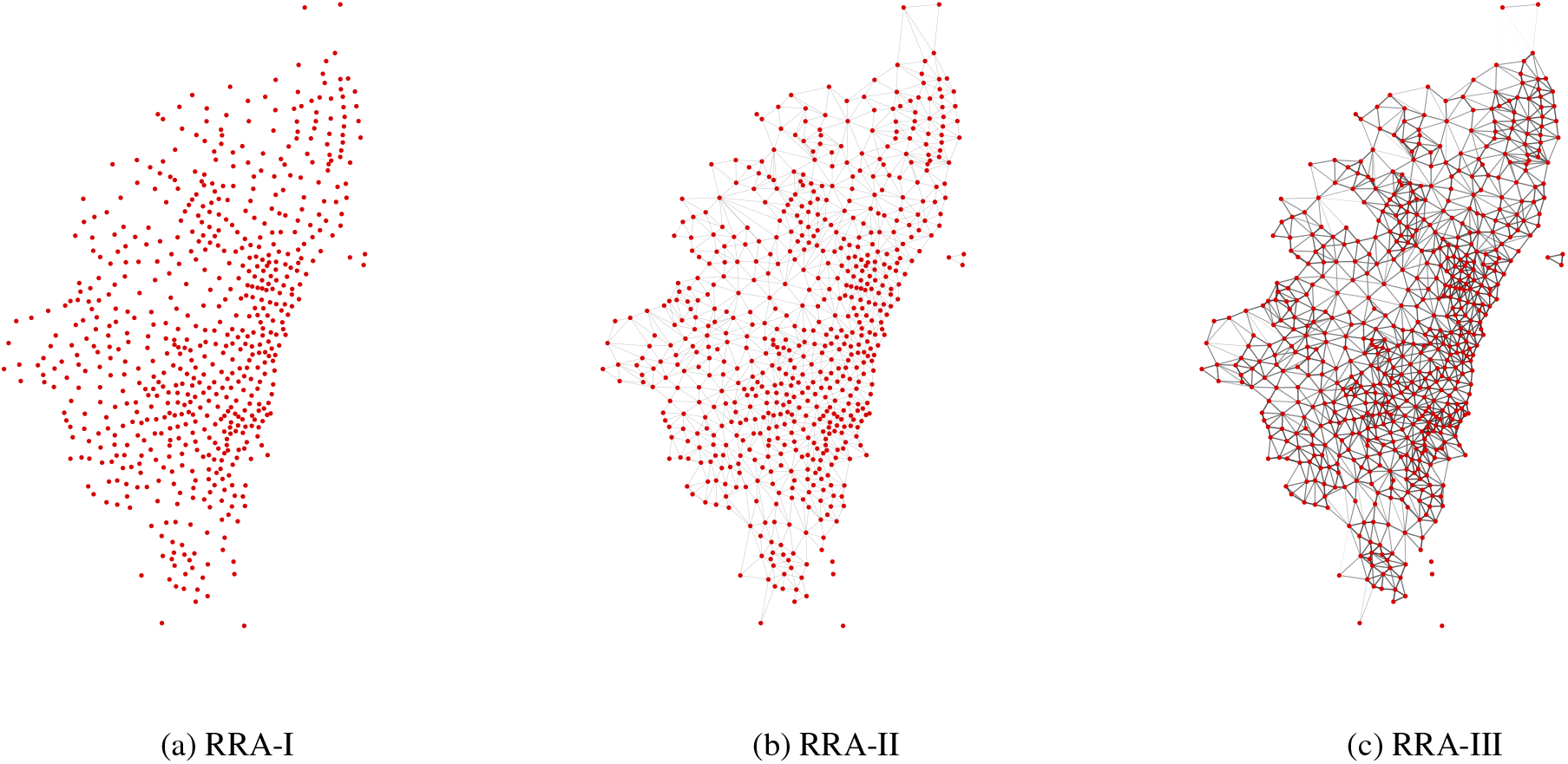
Complex network representation of the three RRA configurations. RRA-I(left), RRA-II (middle), and RRA-III (right).

### 3.2. Risk definition in COVID-19

#### 3.2.1. Pandemic hazard

Natural disasters (i.e., the COVID-19 pandemic) rarely exist, because disasters are social, arising from a combination of hazard and vulnerability, with vulnerability as the causative factor. The disaster occurs at multiple levels simultaneously, with responses to a hazard exposing as many vulnerability problems as the original hazard. The failure to heed to the pandemic plans alongside the lack of health-care accessible to everyone meant that the hazard could not be addressed effectively and vulnerability fundamentals were revealed (Kelman, 2020).

Then, based on mathematical calculations, we assume that the hazard (the new coronavirus) is constant for all spatial units; that is ∀*i* ∈ 𝒮, *H*(*i*) = 1.

#### 3.2.2. Pandemic vulnerability

Three domains are proposed on (Prieto et al., 2021) to deibe the vulnerability for the COVID-19 in Bogotá, Colombia. Those domains are: (i) Where and how he/she lives (life), (ii) Where and how he/she works (work), and (iii) Where and how he/she moves around (movement). Table 1 shows the domains proposed, the vulnerability factors associated with them, and the dataset used to calculate the values for each factor.

**Table 1.**
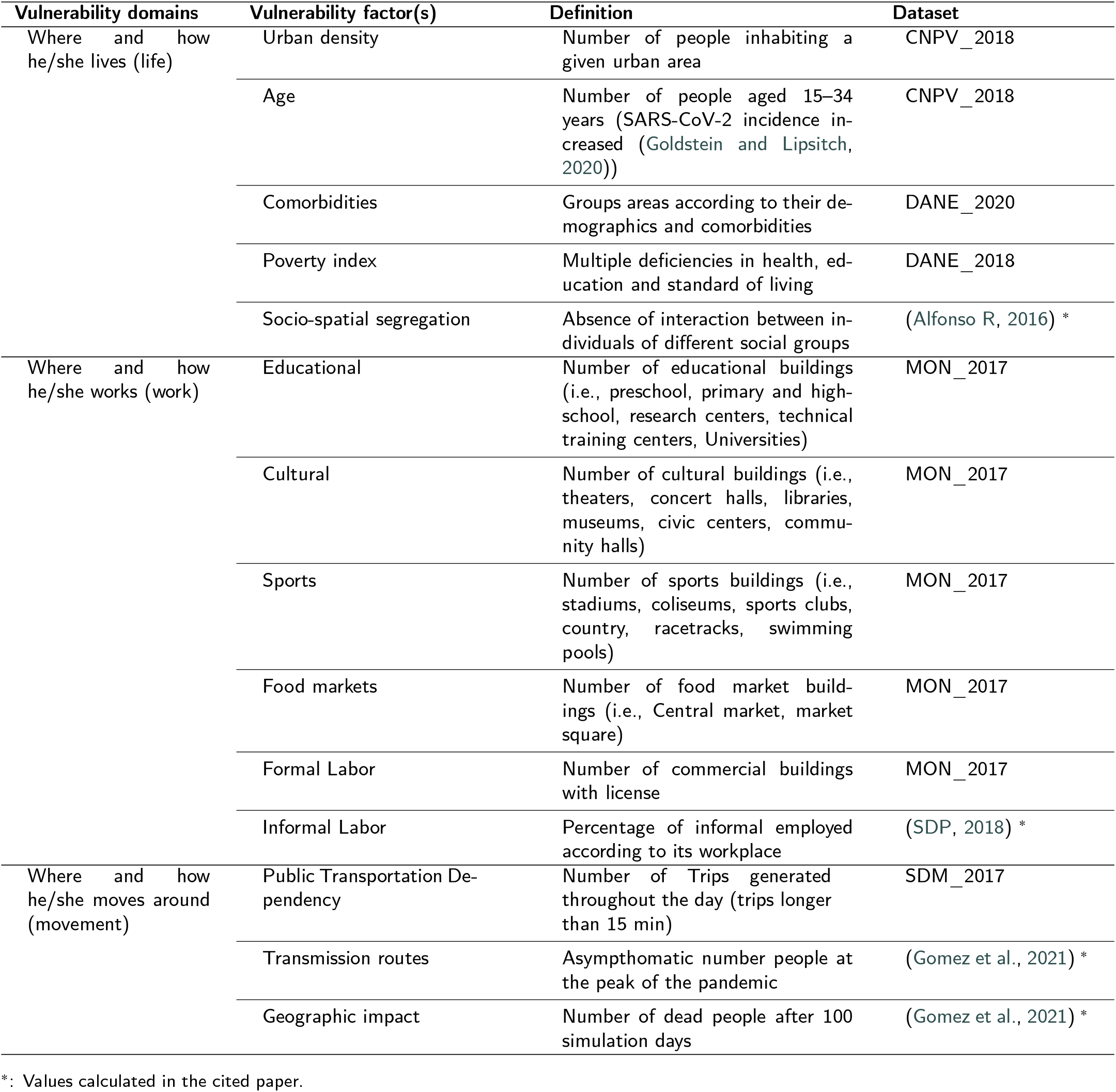
Vulnerability domains for the COVID-19 case in Bogotá, Colombia (Prieto et al., 2021).

- **Where and how he/she lives (life)**: Several demographic factors influence the degree of risk of the geographic areas in a pandemic. The literature emphasizes factors such as *urban density, age*, urban living (i.e., *socio-spatial segregation*), literacy and healthcare quality (i.e., *poverty index*). Further, most data on the COVID-19 pandemic suggest that people with underlying *comorbid* conditions are more vulnerable than people without them.
- **Where and how he/she works (work)**: Urban sectors with high-density facilities (i.e, *educational buildings, cultural buildings, sport buildings, food markets*, all *formal labor, informal labor*) are more vulnerable to the spread of contagious diseases due to space limitations within and between households, growth and mobility, and limited water, sanitation, and hygiene (WASH) infrastructure.
- **Where and how he/she moves around (movement)**: Understanding the *public transportation dependency, transmission routes*, and infection vulnerability factors (i.e., *geographic impact*) provides the baseline for epidemiological modeling that can inform the planning of response and containment efforts to reduce the likelihood of the disease spreading.

Then, based on mathematical calculations, a spatial unit *i* has associated three vulnerable factors: life, work, and movement; that is ∀*i* ∈ 𝒮, *V* (*i*) = {*V*_*life*_(*i*), *V*_*work*_(*i*), *V*_*movement*_(*i*)}.

### 3.3. Risk assessment in COVID-19

Taking the hazard as constant *H*(*i*) = 1, we have only to worry about the Vulnerability Assessment. Previous works (Prieto et al., 2021) proposed a framework for Urban Vulnerability Assessment (UVA) that condense a set of vulnerability factors (Table 1) into a vulnerability index that allowed us to establish and rank potentially vulnerable urban areas in Bogotá. To build the vulnerability index for the three vulnerable domains (life, work, and movement), the following steps are applied:

1. The raw data for each factor is normalized across all spatial units over the range 0 (best) to 1 (worst). The normalization is already calculated in (Prieto et al.,2021).
2. Synthesize the normalized information of all spatial units into *k* partitions which groups spatial units with similar vulnerability profiles. We use *k* = 10 to generate 10 ranges of vulnerabilities (from 0.5 to 0.95, with step of 0.1).
3. The clusters’ centroids of each group are used to sort the vulnerability factors in descending order. This sort is interpreted as vulnerability ranking which is used for the analysis.
4. Then, to aggregate the *L* ranks (one for each vulnerable factor, then for life *L* = 5, work *L* = 6, movement *L* = 3) in a unique vulnerability ranking the Borda’s method is used.
5. The unique vulnerability ranking is then transformed into a vulnerability index, where a higher rank indicates higher vulnerability.

The output of this process is the three vulnerability index (one for each domain). Finally, to quantify the risk we follow the risk definition presented in 1.

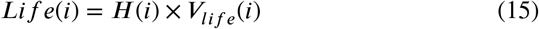

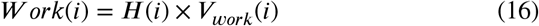

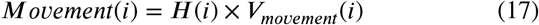

where *Life*(*i*), *W ork*(*i*) and *Movement*(*i*) are the life risk, work risk and movement risk, respectively; for the spatial unit *i*. Fig. 4 shows the final three risk indexes.

**Fig. 4:**
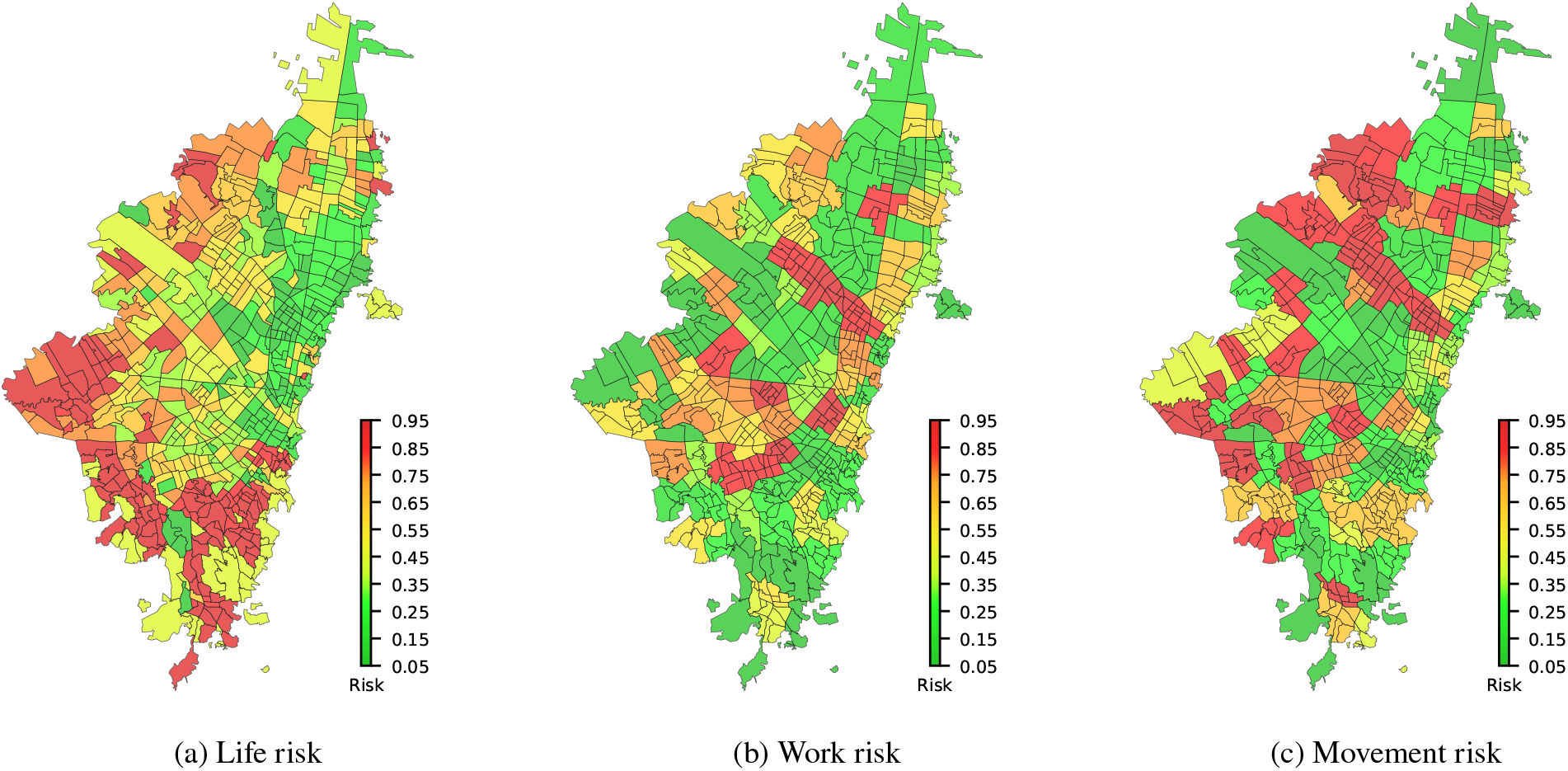
Risk indexes generated for the current COVID-19 pandemic in Bogota, Colombia.

### 3.4. Risk strategy in COVID-19

#### 3.4.1. Experimental Setup

To solve the formulated multi-objective risk-based resource allocation problem, a comparison with different multiobjective algorithms (MOEA/D (Zhang and Li, 2007), NSGA-III (Deb and Jain, 2013), RVEA (Cheng et al., 2016), and ARMOEA (Tian et al., 2017)) was made on different configurations of MoRRA (i.e., RRA-I, RRA-II, RRA-III) and different allocation percentages (i.e., *T* = 10%, *T* = 25%, *T* = 50%). Here, the allocation percentages (amount of resource) *T* is the total of spatial units where a resource should be allocated (i.e., *T* = 10% means that only 10% of the total space units will receive the resource). Also, the impact factor *r*_*i*_ is assumed to 0.5, which means that allocates a resource in the spatial unit *i* would reduce the risk in half.

##### Compared Algorithms

The following four state-of-the-art algorithms for multi-objective functions are considered as peer algorithms.

- MOEA/D (Zhang and Li, 2007): It is representative of the decomposition-based method, the basic idea of MOEA/D is to decompose a MOP into several single-objective optimization subproblems through aggregation functions and simultaneously optimizes them.
- NSGA-III (Deb and Jain, 2013): It is based on decomposition with Pareto-adaptive weight vectors. This approach automatically adjusts the weight vectors by the geometrical characteristics of the Pareto front.
- RVEA (Cheng et al., 2016): It is a scalarization approach, and termed angle penalized distance approach that dynamically adjusts the distribution of the reference vectors to balance convergence and diversity of the solutions in the PFs.
- ARMOEA (Tian et al., 2017): It uses an enhanced inverted generational distance indicator, in which an adaptation method adjusts a set of reference points based on the indicator contributions of candidate solutions.

##### Performance Metrics

To evaluate the performance of different MOEAs for the RRA problems, each algorithm was run for the same number of generations, and the resulting solutions (known as Pareto front approximations), are compared using functions that measure two qualities: (i) solution accuracy, i.e., to determine how similar the solution is to the true Pareto front and (ii) solution diversity, i.e., to evaluate how well distributed are the points in the solution. We selected two of the most used metrics called Δ_*p*_ (Schutze et al., 2012) and Coverage over Pareto Front (*C P F*) (Tian et al., 2019) to compare the accuracy and the diversity of the solutions found by the different algorithms.

##### Parameter Settings

In this subsection, we first present the general parameter settings for the experiments, and afterward, the specific parameter settings for each algorithm in comparison are given.

1. **MoRRA parameters:** We made the experiment with different configurations of RRA (RRA-I, RRA-II, RRA-III) and allocation percentages (*T* = 10%, *T* = 25%, *T* = 50%). The decision variables are equal to the number of urban sectors in Bogotá (*D* = 631) and the objective functions are fixed to 3 (*M* = 3) representing the different risks.
2. **Settings for the operators**: We select the simulated binary crossover (SBX) (Deb et al., 1995) and the polynomial mutation (PM) (Deb and Goyal, 1996) as genetic operators for our experiments. For the SBX, the distribution index is set to *η*_*c*_ = 30 and the crossover probability *p*_*c*_ = 1.0 is used in all algorithms; for PM the distribution index and the mutation probability are set to *η*_*m*_= 20 and *p*_*m*_ = 1/n, respectively.
3. **Population size**: For all algorithms, the population size is determined by the simplex-lattice design factor *H* together with the objective number *M* (Das and Dennis, 1998). Then the population size using this approach is set to 105 individuals.
4. **Termination Condition**: Every algorithm stops when the number of function evaluations reaches the maximum number. For all configurations and allocation percentages the maximal number of generations is set to 10000.
5. **Specific Parameter Settings in Each Algorithm**: For MOEA/D, the weights vectors are calculate using the penalty-based boundary intersection (PBI), the neighborhood size *T* is set to 20, and the penalty parameter *θ* in PBI is set to 5, as recommended in (Zhang and Li, 2007). For RVEA, the parameter controlling the rate of change of penalty (*α*) and the frequency of employing reference vector adaptation (*fr*) are fixed in 2 and 0.1, respectively, as recommend in (Cheng et al., 2016).

#### 3.4.2. Pareto front

The statistical results of the Δ_*p*_ and *C P F* metrics values obtained by the four algorithms for the different configurations and allocation percentages over 20 independent runs are summarized in Table 2, where the best results are highlighted. The Wilcoxon rank-sum test is adopted to compare the results obtained by the four compared algorithms at a significance level of 0.05 (here, the MOEA/D algorithm is taken as the reference’s algorithm). Symbol ‘+’ indicates that MOEA/D is significantly outperformed by the compared algorithm according to a Wilcoxon rank-sum test, while ‘−’ means that the compared algorithm is significantly outperformed by MOEA/D. Finally, ‘≈’ means that there is no statistically significant difference between the results obtained by MOEA/D and the compared algorithm. It can be seen that MOEA/D shows the best overall performance among the four compared algorithms over the Δ_*p*_ metric in the experiments, while RVEA shows the best overall performance over the *C P F* metric in experiments. The results obtained by RVEA (good performance on Δ_*p*_ and *C P F*) in the different configurations and allocation percentages will be used in the rest of the paper for the following results.

**Table 2.**
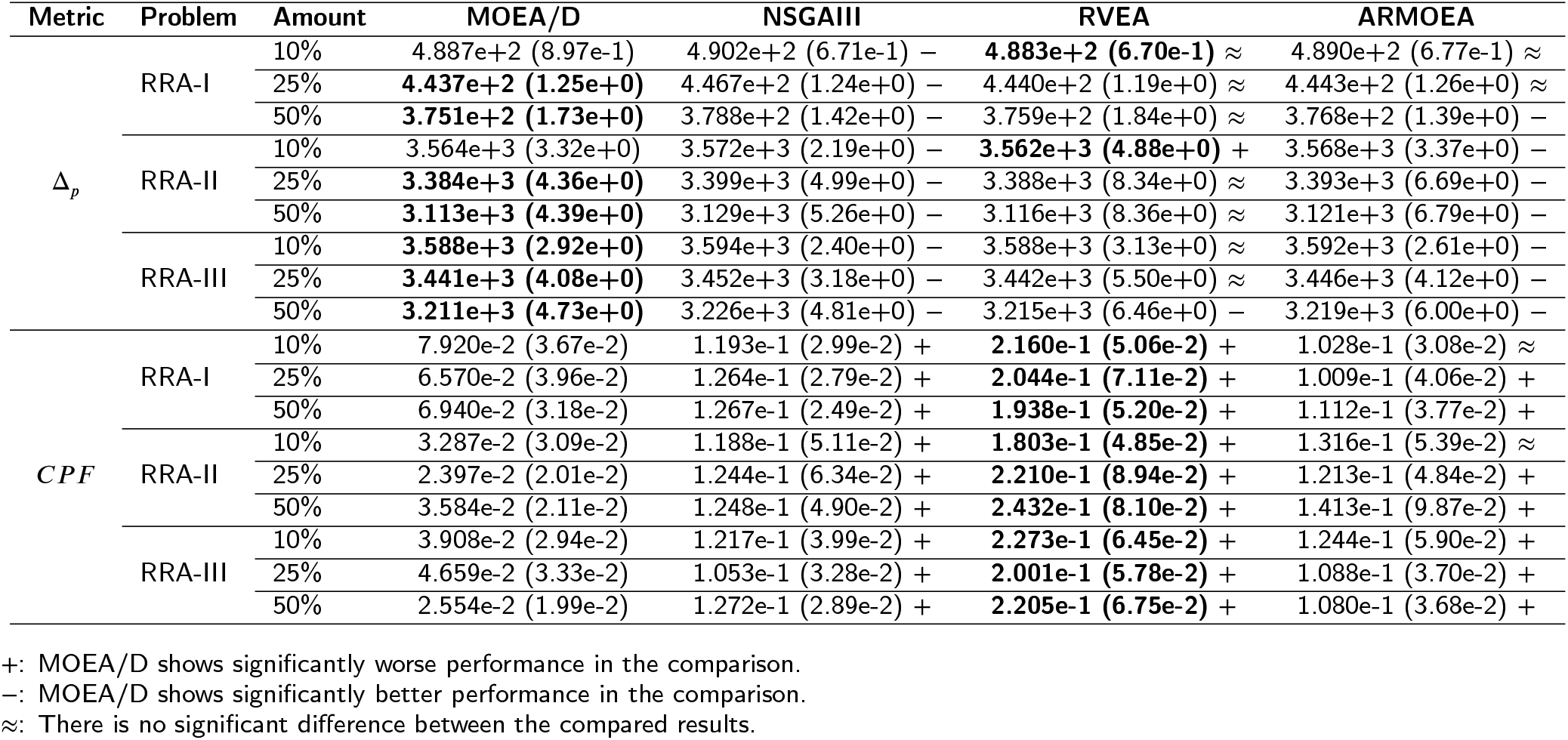
Statistics Δ_*p*_ and *C P F* metric values of the Pareto-optimal solutions founded by the four compared algorithms for the different RRA configurations and amount allocations. The numbers in parentheses are the standard deviations.

The range of the *non-dominated* solutions found with VEA are shown in Fig. 5. The *Pareto front* behavior shows promising convergence performance as well as a good distribution on problems with different configurations and allocation percentages.

**Fig. 5:**
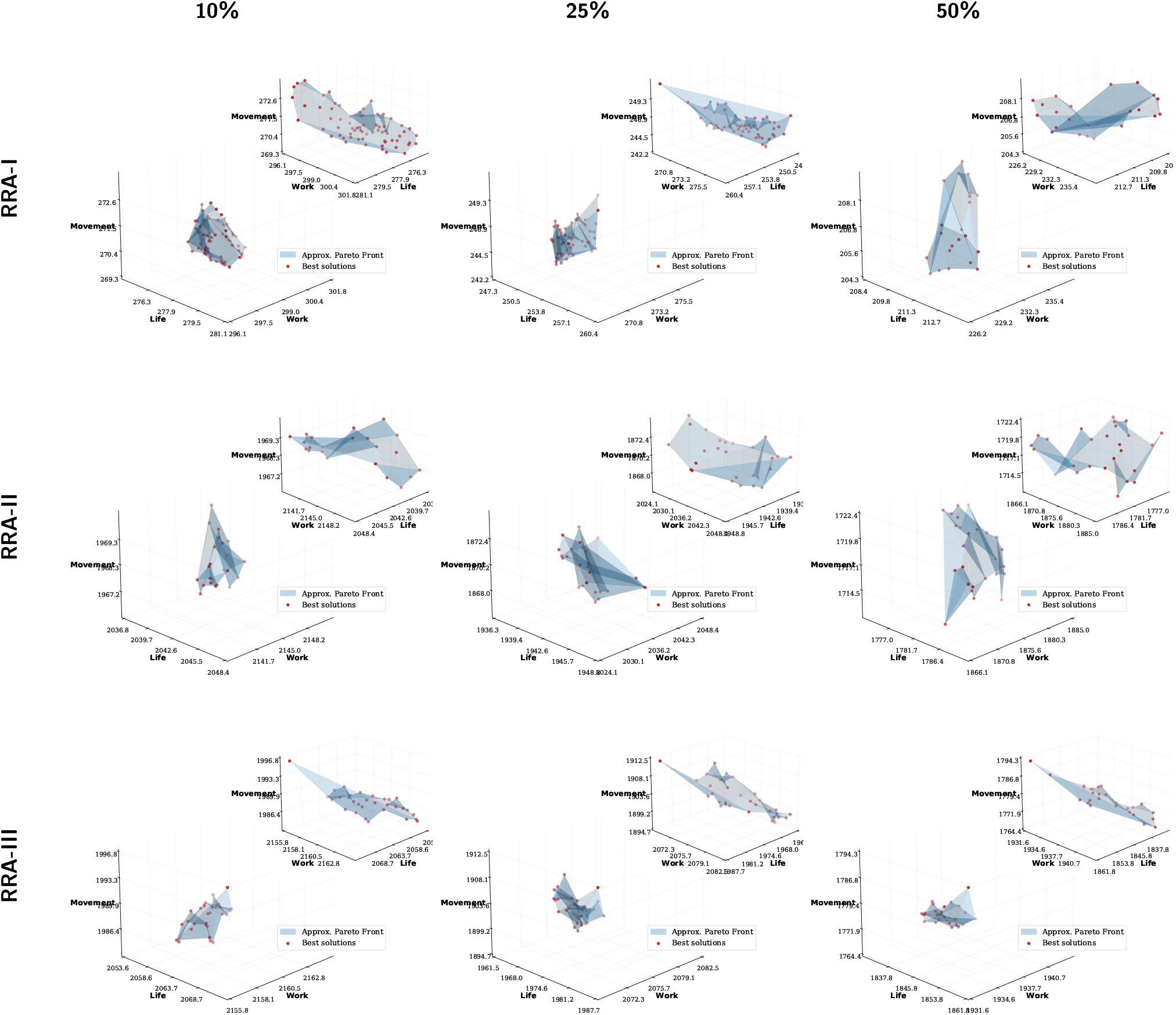
The approximate Pareto optimal solutions obtained by RVEA on problems with different configurations (RRA-I, RRA-II, RRA-III) and allocation percentages (10%, 25%, 50%).

#### 3.4.3. Decision making

To visualize the solution in the Bogotá network map, a pseudo-weight vector approach proposed in (Deb, 2011) is used. This method calculates the normalized distance to the best solution regarding each objective.

First, we select one solution with equal pseudo-weights _*life*_ = 0.33, *w*_*work*_ = 0.33, *w*_*movement*_ = 0.33) for each different RRA configurations and an allocation percentages equal to %10 (Fig 6). The results shows an interesting scenario where the spatial correlation between urban sectors is not remarkable getting an unbiased risk-based resource allocation for COVID-19.

**Fig. 6:**
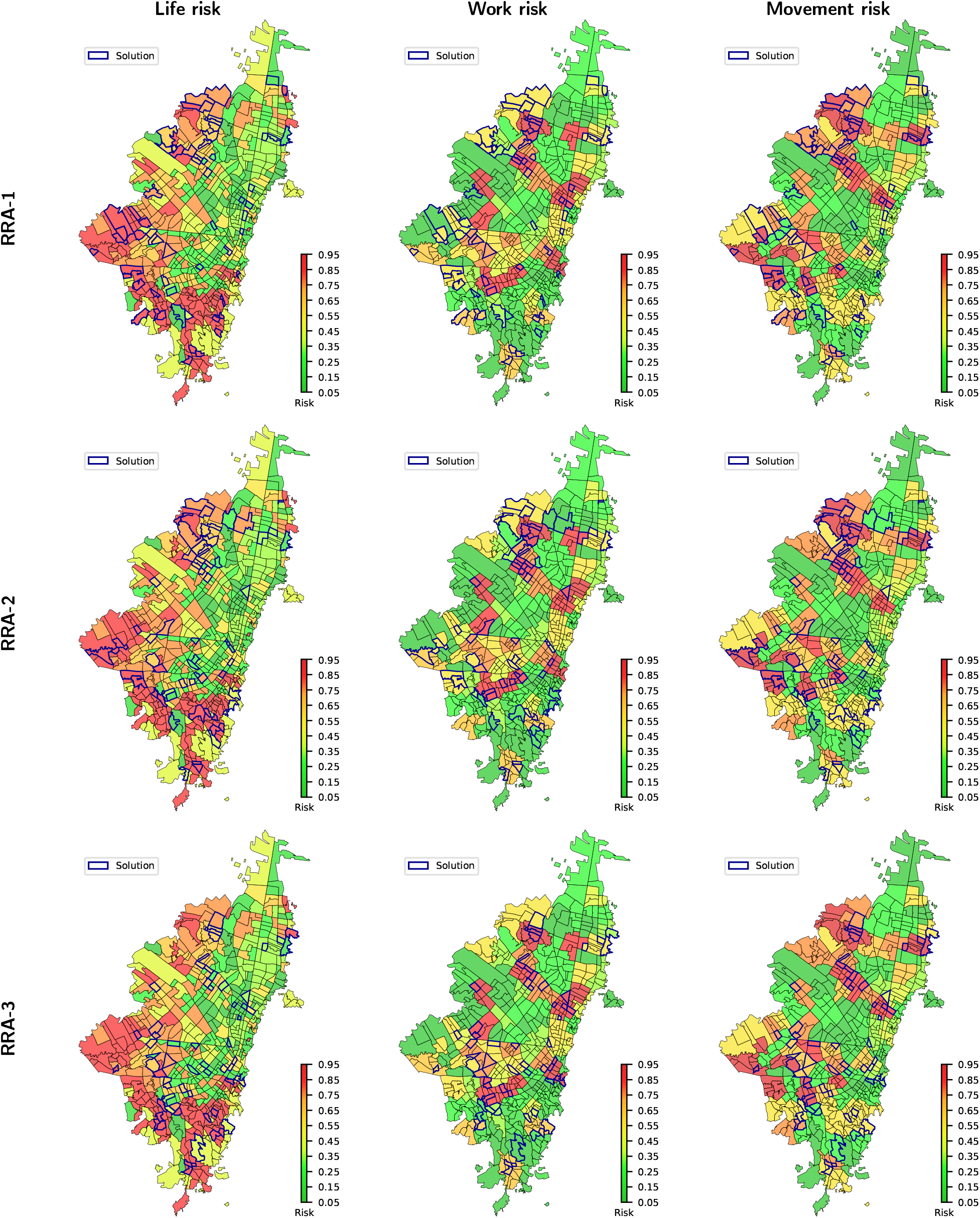
Solution of MoRRA visualized in the Bogotá complex network with 10% of allocation percentage and using equal pseudo-weights (*w*_*life*_ = 0.33, *w*_*work*_ = 0.33, *w*_*movement*_ = 0.33).

Further, in order to support the decision-maker to find the most preferred solution with a good balance between robustness and the nominal quality, different combinations of pseudo-weights are applied in solutions with RRA-III configuration and 10% of allocation percentage (Fig. 7). In the first combination, we give more weight to life risk (*w*_*life*_ = 0.8, *w*_*work*_ = 0.1, *w*_*movement*_ = 0.1). In the second combination, we give more weight to work risk (*w*_*life*_ = 0.1, *w*_*work*_ = 0.8, *w*_*movement*_ = 0.1). Finally, in the third combination, we give more weight to movement risk (*w*_*life*_ = 0.1, *w*_*work*_ = 0.1, *w*_*movement*_ = 0.8). The results indicate that the MoRRA framework proposed could be used to recommend actions for before, during, and after a pandemic that is, to planning and coordination efforts through leadership and coordination across sectors, to assess if the risk of a pandemic could increase in specific geographic areas.

**Fig. 7:**
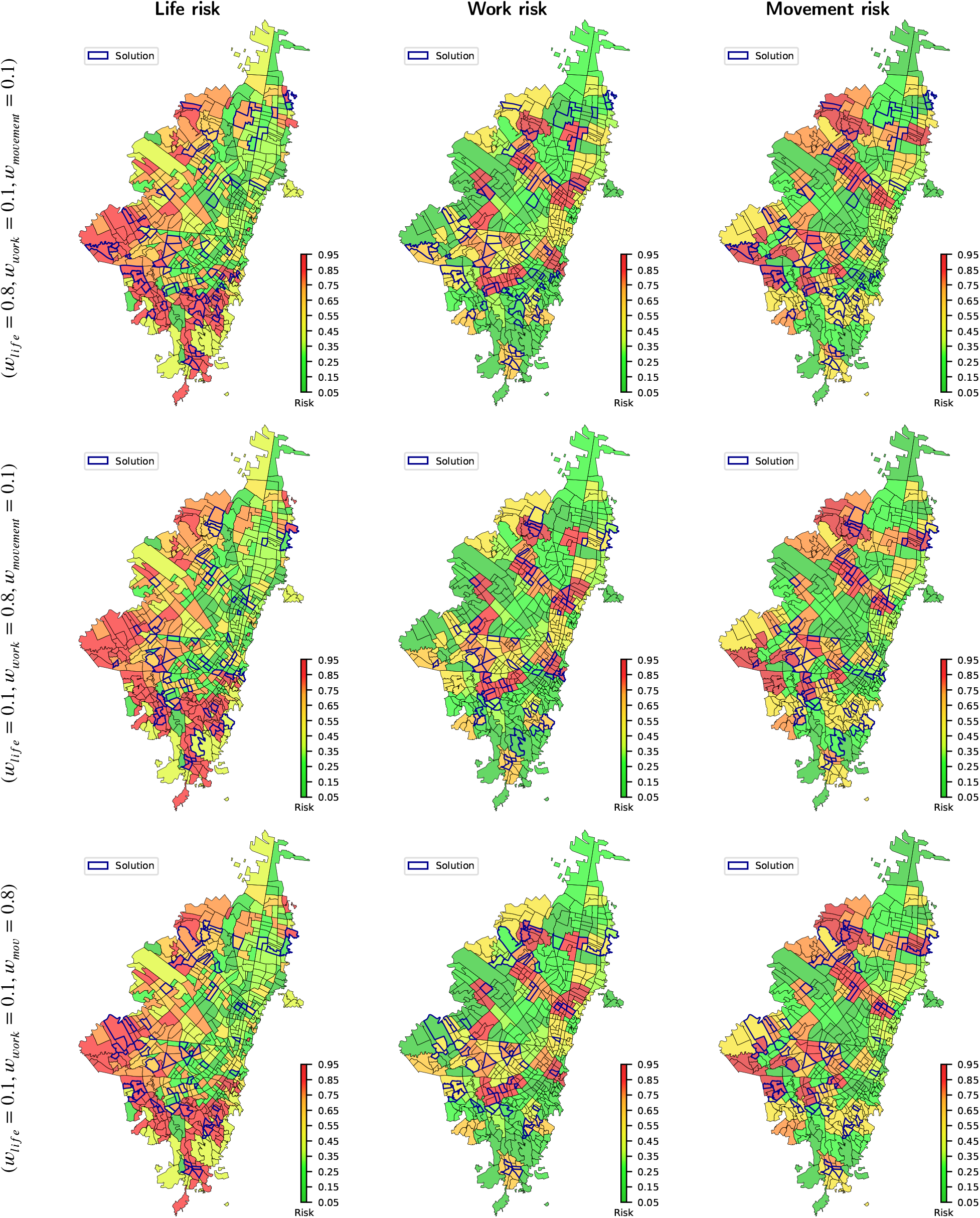
Solution of MoRRA visualized in the Bogotá complex network with different configurations of RRA and 10% of allocation percentage and using different combinations of pseudo-weights.

## 4. Conclusions and Future Work

A Multi-objective Risk-based Resource Allocation (MoRRA) framework for Pandemic Preparedness is proposed. MoRRA could be used to build evidence for planning, modeling, and epidemiological studies to better inform the public, policymakers, and international organizations and funders as to where and how to improve surveillance, response efforts, and delivery of resources. The proposed MoRRA is tested in the current COVID-19 Pandemic in Bogotá city, the largest and crowded city in Colombia. MoRRA creates not only one, but a set of risk indices (i.e., life, work, movement) and uses them to apply the risk-based resource allocation.

Although the risk factors involved in the framework are structural, the proposed approach is flexible, does not require expert support or knowledge, and allows policy-makers, and international organizations to prioritize resource allocation in short and long-term actions for affected populations in a city. For instance, using the solutions with more weight in the movement risk a short-actions (i.e, staying home, limit close contact, avoid crowds, limit non-essential travel) can be taken to reduce the risk in the city. Further, using the solutions with more weight in the life risk, it is possible to advance in long-term territorial reorganization plans (i.e., reduce socio-spatial segregation, decent housing, bio-secure protocols for high-density facilities) as our results indicate for the COVID-19 in the urban area of Bogotá.

### A. Matrix Notation of RRA

To derive (9) for (4), the Euclidean Complex Network *G* is represented with its adjacency matrix *A*. Then, we want to demonstrate.

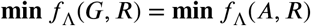

when *f*_Λ_(*A, R*) is written as.

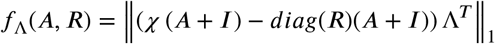

Here, *χ* is the Indicator function that determines when a value of *A* is different to 0, *I* is the identity matrix, *diag* is the function that diagonalizes the vector of resources *R*, and A are the risks associated with the demand points.

Expanding *f*_Λ_(*A, R*), we have.

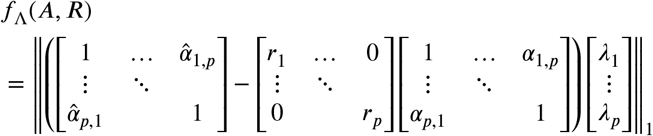

Solving.

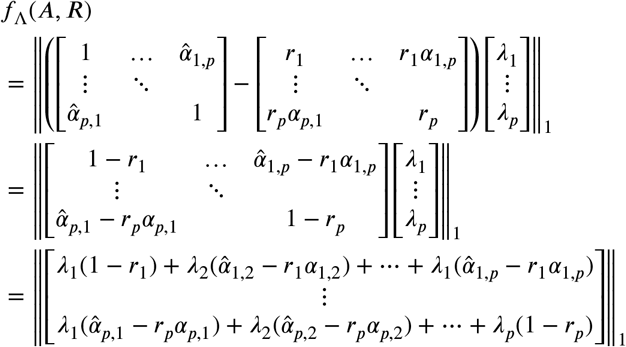

Grouping similar terms.

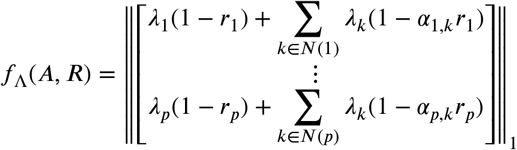

Each term in the column vector can be written as (3).

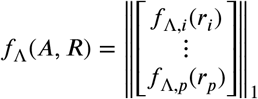

Applying 1-Norm.

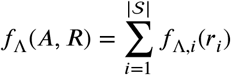

So, it is sufficient to prove that (4) could be written as (9) when the graph *G* is representing by its adjacency matrix *A*.

## Data Availability

No medical data from external sources is used in this paper. Demographic data of Bogotá city is available on Colombian government sites.

## Notes

### Competing Interest Statement

The authors have declared no competing interest.

### Funding Statement

No additional funding was granted for writing this paper.

